# Comparative effectiveness of Paxlovid versus sotrovimab and molnupiravir for preventing severe COVID-19 outcomes in non-hospitalised patients: observational cohort study using the OpenSAFELY platform

**DOI:** 10.1101/2023.01.20.23284849

**Authors:** Bang Zheng, John Tazare, Linda Nab, Amir Mehrkar, Brian MacKenna, Ben Goldacre, Ian J Douglas, Laurie A Tomlinson, The OpenSAFELY Collaborative

## Abstract

**Objective:** To compare the effectiveness of Paxlovid vs. sotrovimab and molnupiravir in preventing severe COVID-19 outcomes in non-hospitalised high-risk COVID-19 adult patients.

**Design:** With the approval of NHS England, we conducted a real-world cohort study using the OpenSAFELY-TPP platform.

**Setting:** Patient-level electronic health record data were obtained from 24 million people registered with a general practice in England that uses TPP software. The primary care data were securely linked with data on COVID-19 infection and therapeutics, hospital admission, and death within the OpenSAFELY-TPP platform, covering a period where both Paxlovid and sotrovimab were first-line treatment options in community settings.

**Participants:** Non-hospitalised adult COVID-19 patients at high risk of severe outcomes treated with Paxlovid, sotrovimab or molnupiravir between February 11, 2022 and October 1, 2022.

**Interventions:** Paxlovid, sotrovimab or molnupiravir administered in the community by COVID-19 Medicine Delivery Units.

**Main outcome measure:** COVID-19 related hospitalisation or COVID-19 related death within 28 days after treatment initiation.

**Results:** A total of 7683 eligible patients treated with Paxlovid (n=4836) and sotrovimab (n=2847) were included in the main analysis. The mean age was 54.3 (SD=14.9) years; 64% were female, 93% White and 93% had three or more COVID-19 vaccinations. Within 28 days after treatment initiation, 52 (0.68%) COVID-19 related hospitalisations/deaths were observed (33 (0.68%) treated with Paxlovid and 19 (0.67%) with sotrovimab). Cox proportional hazards model stratified by region showed that after adjusting for demographics, high-risk cohort categories, vaccination status, calendar time, body mass index and other comorbidities, treatment with Paxlovid was associated with a similar risk of outcome event as treatment with sotrovimab (HR=1.14, 95% CI: 0.62 to 2.08; P=0.673). Results from propensity score weighted Cox model also showed comparable risks in these two treatment groups (HR=0.88, 95% CI: 0.45 to 1.71; P=0.700). An exploratory analysis comparing Paxlovid users with 802 molnupiravir users (11 (1.37%) COVID-19 related hospitalisations/deaths) showed some evidence in favour of Paxlovid but with variation in the effect estimates between models (HR ranging from 0.26 to 0.61).

**Conclusion:** In routine care of non-hospitalised high-risk adult patients with COVID-19 in England, no substantial difference in the risk of severe COVID-19 outcomes was observed between those who received Paxlovid and sotrovimab between February and October 2022, when different subvariants of Omicron were dominant.

## Background

On December 16, 2021, COVID-19 Medicine Delivery Units (CMDUs) were launched across England to provide antiviral medicines and neutralising monoclonal antibodies (nMAbs) to treat symptomatic COVID-19 patients in community settings who were at high risk of severe outcomes. The clinical guideline from NHS England [1,2] has been revised over time based on emerging evidence and the approval of new medications by the UK Medicines and Healthcare products Regulatory Agency (MHRA). After February 10, 2022, nirmatrelvir plus ritonavir (Paxlovid, an oral antiviral) and sotrovimab (an intravenous nMAb) were both recommended as first-line treatments for non-hospitalised high-risk COVID-19 patients to prevent disease progression [2].

The approval and early routine clinical use of these two medications were mainly supported by evidence from two RCTs in unvaccinated population before the Omicron wave [3,4], with certain vulnerable clinical subgroups being underrepresented (such as immunosuppressed individuals). Several real-world observational studies in non-hospitalised COVID-19 patients during the Omicron era have shown effectiveness of Paxlovid compared with non-users [5,6], but those studies were likely to suffer from confounding by indication, given the large differences in the characteristics and clinical conditions between treated and untreated patients (e.g., untreated patients could have been either of low-risk of severe outcomes, or high-risk but with contraindications to Paxlovid). Comparative effectiveness analysis for treatments prescribed under similar clinical indications, with careful consideration of the contraindications and drug interactions for the use of Paxlovid [2], would tend to ensure comparability of participants, leading to more robust findings. Comparative evidence is also more informative for a clinical setting where the decision is over which treatment to use rather than whether to treat or not.

The latest WHO guideline makes a strong recommendation against sotrovimab for the treatment of non-severe COVID-19 patients due to its reduced activity against Omicron variants in in vitro studies [7]. In the updated version of NHS England guideline for non-hospitalised COVID-19 patients published on November 28, 2022, sotrovimab was de-prioritised and is only to be considered by exception where the available antivirals are contraindicated or unsuitable [8]. In contrast, Paxlovid remains as the first-line option in both guidelines [7,8]. Validation research directly comparing the effectiveness of sotrovimab and Paxlovid in preventing severe outcomes in routine usage is thus urgently needed for the development of an evidence-based clinical management pathway.

Therefore, following the national CMDU rollout of these two first-line medications since February 10, 2022, this real-world observational study aimed to compare the effectiveness of Paxlovid and sotrovimab on preventing severe outcomes in non-hospitalised high-risk COVID-19 patients across England, utilising the near real-time electronic health record (EHR) data in the OpenSAFELY-TPP platform [9]. As an exploratory analysis, we also compared the risks of severe outcomes between Paxlovid users and those treated with molnupiravir [10] (a third-line antiviral in NHS England guideline) during the study period, with the assumption that this group is more comparable to Paxlovid users than untreated controls.

## Methods

### Data Source

Primary care records managed by the GP software provider, TPP were linked to the Office for National Statistics (ONS) death data through OpenSAFELY, a data analytics platform created by our team on behalf of NHS England to address urgent COVID-19 research questions (https://opensafely.org). OpenSAFELY provides a secure software interface allowing the analysis of pseudonymised primary care patient records from England in near real-time within the EHR vendor’s highly secure data centre, avoiding the need for large volumes of potentially disclosive pseudonymised patient data to be transferred off-site. This, in addition to other technical and organisational controls, minimises any risk of re-identification. Similarly pseudonymised datasets from other data providers are securely provided to the EHR vendor and linked to the primary care data. Further details on our information governance can be found on [information governance and ethics](https://docs.google.com/document/d/1ZdaeKj8DXnwWoxK5HuQhkBEHahdrcx3boq4sobwAVsA/edit#heading=h.dyd1pe1wp9sk).

The dataset analysed within OpenSAFELY is based on 24 million people currently registered with GP surgeries using TPP SystmOne software. It includes pseudonymised data such as coded diagnoses, medications and physiological parameters. Patient-level vaccination status is available in the GP records directly via the National Immunisation Management System (NIMS). No free text data are included. The following linked data were also used for this study: accident and emergency (A&E) attendance and in-patient hospital spell records via Secondary Uses Service (SUS); national coronavirus testing records via the Second Generation Surveillance System (SGSS); and the “COVID-19 therapeutics dataset”, a patient-level dataset on antiviral and nMAbs treatments, newly sourced from NHS England, derived from Blueteq software that CMDUs use to notify NHS England of COVID-19 treatments [9].

### Study Design and Population

We conducted a population-based cohort study with all adults (≥18 years old) within OpenSAFELY-TPP who had treatment records of either Paxlovid, sotrovimab or molnupiravir from CMDUs between February 11, 2022 and October 1, 2022. During this treatment period, there was relative clinical equipoise between Paxlovid and sotrovimab (both as first-line options in NHS England guideline) [2]. In addition, eligible patients in this study were required to be non-hospitalised for COVID-19 at treatment initiation (as recorded in COVID-19 therapeutics dataset [9]), and be registered in GP surgeries before treatment. Patients were excluded if they had treatment records of any other antivirals or nMAbs for COVID-19 before receiving the treatment under investigation (n=76). Patients with treatment records of other antivirals or nMAbs after receiving the treatment under investigation were censored at the start date of the second treatment (n=31).

According to the eligibility criteria from NHS England [2], to have received an nMAb or antiviral treatment for COVID-19 in the community during this period, patients needed to have SARS-CoV-2 infection confirmed by polymerase chain reaction (PCR) testing or lateral flow test, onset of COVID-19 symptoms within the last five days, have no signs of recovery and not require hospitalisation for COVID-19 or supplemental oxygen specifically for the management of COVID-19 symptoms before treatment, and be a member of at least one of the following ten high-risk cohorts: patients with Down syndrome, a solid cancer, a haematological disease or stem cell transplant, renal disease, liver disease, immune-mediated inflammatory disorders, primary immune deficiencies, HIV/AIDS, solid organ transplant, or rare neurological conditions.

To be noted, there was caution about treatment with Paxlovid if patients had a history of advanced decompensated liver cirrhosis, stage 3-5 chronic kidney disease (or stage 4-5 in the updated guideline), solid organ transplant, or were taking any of the medications with potential drug-drug interactions listed in the NHS England guideline [2]. Therefore, we detected and excluded all patients with these restrictions based on medication records (prescription of any contraindicated medications in “Drugs do not use” and “Drugs consider risks and benefits” codelists [11] within 180 days before treatment), diagnosis codes and clinical tests (eGFR or creatinine) in OpenSAFELY-TPP to ensure comparability between participants treated with Paxlovid and sotrovimab or molnupiravir (**Figure 1**).

**Figure 1.**
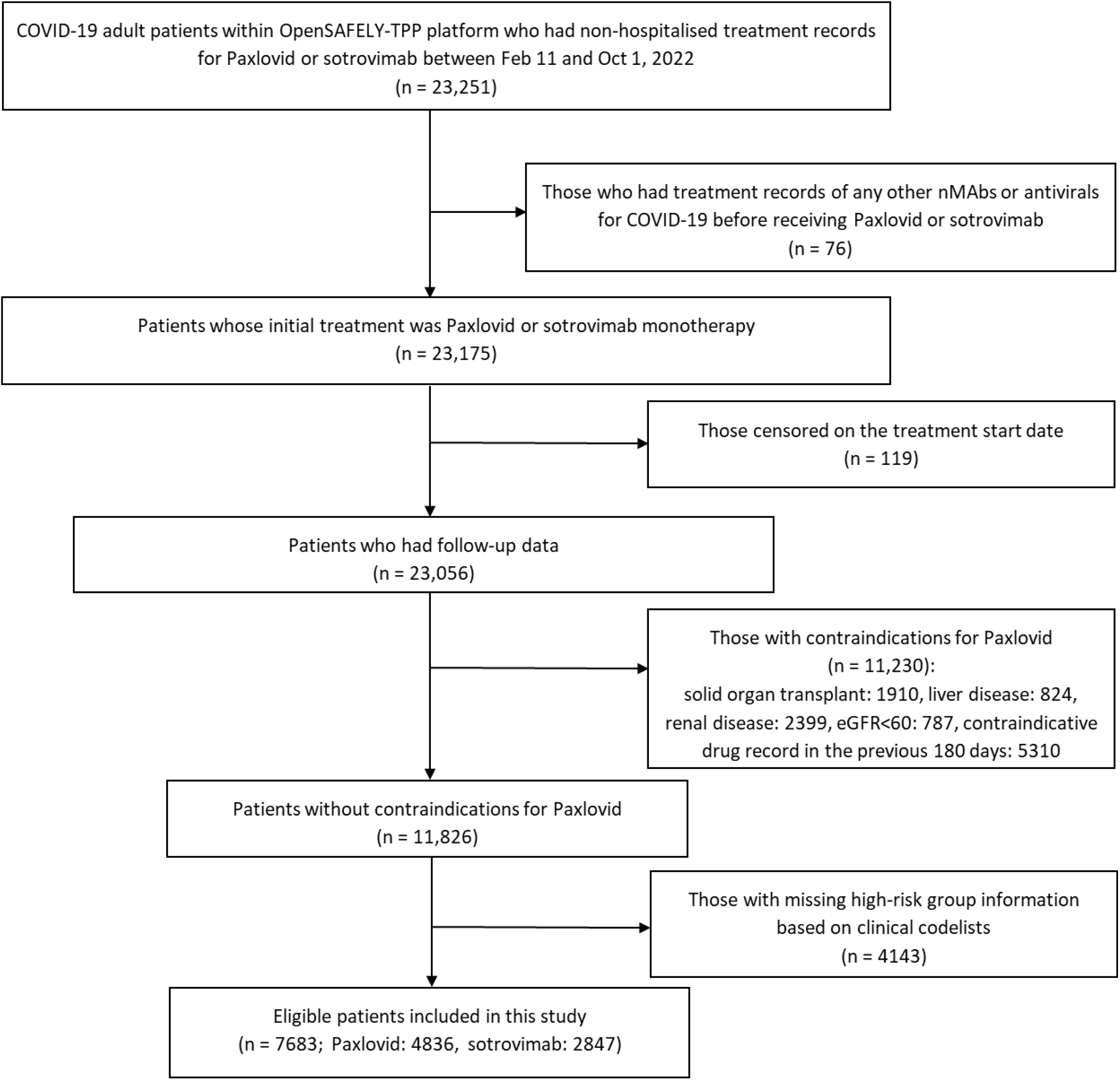
Flowchart for study participants.

### Study Measures

#### Exposure

The exposure of interest was treatment with Paxlovid versus sotrovimab administered by CMDUs in the main study, and treatment with Paxlovid versus molnupiravir in the exploratory analysis. Exposure status and date of treatment of each patient were ascertained from the COVID-19 therapeutics dataset.

#### Outcome

The primary outcome was COVID-19 related hospitalisation (based on primary diagnosis ascertained from SUS) or COVID-19 related death (based on underlying/contributing causes of death from ONS) within 28 days after treatment initiation. Secondary outcomes were 28-day all-cause hospital admission or death, and 60-day COVID-19 related hospitalisation/death. To exclude events where patients were admitted in order to receive sotrovimab or other planned/regular treatment, we did not count admissions coded as “elective day case admission” or “regular admission” in SUS or day cases detected by the same admission and discharge dates as hospitalisation events.

#### Covariates

The following potential confounding factors were extracted at baseline: age, sex, NHS region, ethnicity, Index of Multiple Deprivation (IMD, as quintiles derived from the patient’s postcode at lower super output area level), rural-urban classification (derived from patient’s postcode), calendar time (to account for secular trend of prescription and COVID-19 outcomes), COVID-19 vaccination status (unvaccinated, one vaccination, two vaccinations, three vaccinations, or four or more), positive test date for SARS-CoV-2 infection (PCR or lateral flow test), BMI (most recent record within 10 years), high-risk cohort categories as mentioned above (allowing multiple categories per patient), other comorbidities (diabetes, hypertension, chronic heart diseases, chronic respiratory diseases, learning disabilities, severe mental illness), and care home residency and housebound status. Individuals with missing ethnicity, IMD, BMI or positive SARS-CoV-2 test information were included as “Unknown” category.

### Statistical Analysis

For the comparative effectiveness analysis of Paxlovid vs. sotrovimab, distributions of baseline characteristics were compared between patients in these two treatment groups. Follow-up time of individual patients was calculated from the date of the treatment initiation record, until the date of outcome event, 28 days after treatment initiation, initiation of a second nMAb/antiviral treatment, death, patient de-registration date, or the study end date (December 22, 2022), whichever occurred first.

Risks of 28-day COVID-19 related hospitalisation/death were compared between the two drug groups using Cox proportional hazards models, with time since treatment as the time scale. The Cox models were stratified by region to account for geographic heterogeneity in baseline hazards, with sequential adjustment for other baseline covariates. Model 1 was adjusted for age and sex; Model 2 additionally adjusted for high-risk cohort categories (Down syndrome, solid cancer, haematological disease, immune-mediated inflammatory disorders, immunosuppression, rare neurological conditions); Model 3 further adjusted for ethnicity (White or non-White), IMD quintiles, vaccination status, calendar date (with restricted cubic splines to account for non-linear effect); and Model 4 additionally adjusted for BMI category (<25 kg/m^2^, 25 - <30 kg/m^2^, ≥30 kg/m^2^), diabetes, hypertension, chronic cardiac and respiratory diseases. Missing values of covariates were treated as separate categories. The proportional hazards assumption was assessed by testing for a zero slope in the scaled Schoenfeld residuals for each Cox model.

As an alternative approach to account for confounding bias, we used the propensity score weighting (PSW) method to balance the distributions of relevant covariates between groups. The propensity score (PS) for each patient is defined as the conditional probability of being treated with Paxlovid, estimated with a binary logistic regression of the actual treatment on relevant baseline covariates. The average treatment effect (ATE) weighting scheme based on propensity scores (with and without trimming: approaches discussed further in results) [12,13]) was then applied to the Cox model. Balance check of baseline covariates after weighting was conducted using standardised mean difference (SMD) between groups. Robust variance estimators were used in the weighted Cox model. Similar analyses were conducted for secondary outcomes.

Further exploratory analyses were conducted by different subgroups, including time period with different dominant variants (February 11-May 31 for BA.2, June 1-October 1 for BA.5 [14]), each high-risk cohort, presence of obesity (≥30 vs. <30 kg/m^2^), diabetes, hypertension, chronic cardiac diseases or chronic respiratory diseases, days between test positive and treatment initiation (<3 vs. 3-5 days), age group (<60 vs. ≥60 years), sex and ethnicity (White vs. non-White). Effect modification by each covariate was tested by adding the corresponding interaction term in the stratified Cox model, with Bonferroni correction for multiple testing.

Several sensitivity analyses based on the stratified Cox model were conducted to assess the robustness of main findings, including (1) using complete-case analysis or Multiple Imputation by Chained Equations to deal with missing values in covariates; (2) additionally adjusting for time between test positive and treatment initiation, and time between last vaccination date and treatment initiation; (3) additionally adjusting for rural-urban classification, and other comorbidities and factors that might have influenced clinician’s choice of therapy through the patient’s ability to travel to hospital for an infusion (learning disabilities, severe mental illness, care home residency or housebound status); (4) using restricted cubic splines for age to further control for potential non-linear age effect; (5) excluding patients with treatment records of both sotrovimab and Paxlovid, or any other treatments (i.e., casirivimab, molnupiravir, or remdesivir); (6) excluding patients who did not have a positive SARS-CoV-2 test record before treatment or initiated treatment after 5 days since positive SARS-CoV-2 test; (7) creating a 1-day or 2-day lag in the follow-up start date to account for potential delays in drug administration; (8) excluding those with prescription record of any contraindicative medication for Paxlovid within 1 year before treatment of Paxlovid of sotrovimab, or within 90 days before treatment of Paxlovid of sotrovimab; (9) not excluding those with contraindications or contraindicative medications for Paxlovid, adjusting for these conditions as covariates instead.

Finally, we adopted similar analytical approaches for the exploratory analysis comparing Paxlovid users with molnupiravir users.

### Software and Reproducibility

Data management was performed using Python, with analysis carried out using Stata 16.1. Code for data management and analysis, as well as codelists, are archived online (https://github.com/opensafely/Paxlovid-and-sotrovimab). All iterations of the pre-specified study protocol are archived with version control (https://github.com/opensafely/Paxlovid-and-sotrovimab/tree/main/docs).

## Results

### Patient characteristics

Between February 11 and October 1, 2022, a total of 7683 non-hospitalised COVID-19 patients treated with Paxlovid (n=4836) or sotrovimab (n=2847) were included in the main analysis. The mean age of these patients was 54.3 (SD=14.9) years; 64% were female, 93% White and 93% had three or more COVID-19 vaccinations. Compared with the sotrovimab group, those receiving Paxlovid were slightly younger (53.5 vs. 55.7 years), had a higher proportion of Down syndrome, immune-mediated inflammatory disorders and rare neurological conditions, and lower proportion of solid cancer, haematological disease, diabetes, hypertension, chronic heart diseases and chronic respiratory diseases (**Table 1**). There were also some geographic variations in the prescription of these two drugs and greater use of sotrovimab earlier during the study period. Other baseline characteristics were similar between the two groups (**Table 1**).

**Table 1.** Baseline characteristics of patients receiving Paxlovid or sotrovimab.

| Characteristics | Paxlovid group | Sotrovimab group | Total |
| --- | --- | --- | --- |
| **N** | 4836 | 2847 | 7683 |
| **Age (year), mean (SD)** | 53.5 (14.8) | 55.7 (15.2) | 54.3 (14.9) |
| **Female, n (%)** | 3092 (63.9) | 1822 (64.0) | 4914 (64.0) |
| **White, n (%)** | 4422 (93.0) | 2623 (93.3) | 7045 (93.1) |
| **IMD quintile, n (%)** |  |  |  |
| **1 (most deprived)** | 467 (10.0) | 276 (10.0) | 743 (10.0) |
| **2** | 723 (15.4) | 465 (16.8) | 1188 (15.9) |
| **3** | 1048 (22.3) | 684 (24.7) | 1732 (23.2) |
| **4** | 1195 (25.5) | 682 (24.7) | 1877 (25.2) |
| **5 (least deprived)** | 1262 (26.9) | 658 (23.8) | 1920 (25.7) |
| **Region (NHS), n (%)** |  |  |  |
| **East** | 1180 (24.4) | 860 (30.2) | 2040 (26.6) |
| **London** | 219 (4.5) | 198 (7.0) | 417 (5.4) |
| **East Midlands** | 1129 (23.4) | 643 (22.6) | 1772 (23.1) |
| **West Midlands** | 61 (1.3) | 143 (5.0) | 204 (2.7) |
| **North East** | 125 (2.6) | 121 (4.3) | 246 (3.2) |
| **North West** | 487 (10.1) | 213 (7.5) | 700 (9.1) |
| **South East** | 474 (9.8) | 114 (4.0) | 588 (7.7) |
| **South West** | 535 (11.1) | 413 (14.5) | 948 (12.3) |
| **Yorkshire** | 626 (12.9) | 142 (5.0) | 768 (10.0) |
| **High risk cohorts, n (%)** |  |  |  |
| **Down syndrome** | 204 (4.2) | 71 (2.5) | 275 (3.6) |
| **Solid cancer** | 658 (13.6) | 508 (17.8) | 1166 (15.2) |
| **Haematological disease** | 729 (15.1) | 639 (22.4) | 1368 (17.8) |
| **Immune-mediated inflammatory diseases** | 1846 (38.2) | 973 (34.2) | 2819 (36.7) |
| **Immunosuppression** | 525 (10.9) | 290 (10.2) | 815 (10.6) |
| HIV/AIDS | 14 (0.3) | 7 (0.3) | 21 (0.3) |
| Rare neurological disease | 1290 (26.7) | 628 (22.1) | 1918 (25.0) |
| BMI (kg/m<sup>2</sup>), mean (SD) | 28.1 (6.5) | 28.1 (6.3) | 28.1 (6.4) |
| Comorbidities, n (%) |  |  |  |
| Diabetes | 603 (12.5) | 425 (14.9) | 1028 (13.4) |
| Chronic cardiac disease | 271 (5.6) | 231 (8.1) | 502 (6.5) |
| Hypertension | 1121 (23.2) | 884 (31.1) | 2005 (26.1) |
| Chronic respiratory disease | 729 (15.1) | 498 (17.5) | 1227 (16.0) |
| Vaccination status, n (%) |  |  |  |
| None | 74 (1.5) | 37 (1.3) | 111 (1.4) |
| One vaccination | 74 (1.5) | 26 (0.9) | 100 (1.3) |
| Two vaccinations | 192 (4.0) | 115 (4.0) | 307 (4.0) |
| Three vaccinations | 2449 (50.6) | 1411 (49.6) | 3860 (50.2) |
| Four or more | 2047 (42.3) | 1258 (44.2) | 3305 (43.0) |
| Days between test positive and treatment, median (IQR) | 1 (1-2) | 2 (1-3) | 2 (1-2) |
| Weeks between campaign start and treatment, median (IQR) | 23 (16-30) | 17 (14-28) | 20 (15-30) |

Note: IMD, BMI, ethnicity and positive test date had 223, 830, 119 and 968 missing values, respectively.

### Comparative effectiveness of Paxlovid vs. sotrovimab for the outcome events

During the 28 days of follow-up after treatment initiation, 52 cases (0.68%) of COVID-19 related hospitalisations/deaths were observed, with 33 (0.68%) in the Paxlovid group and 19 (0.67%) in the sotrovimab group. The number of COVID-19 related deaths were 8 in the Paxlovid group and ≤5 in the sotrovimab group.

Results of stratified Cox regression showed that, after adjusting for demographic variables, high-risk cohort categories, vaccination status, calendar date, BMI category and other comorbidities, treatment with Paxlovid was associated with a similar risk of 28-day COVID-19 related hospitalisation/death as treatment with sotrovimab (hazard ratio, HR=1.14, 95% CI: 0.62 to 2.08; P=0.673, with sotrovimab as baseline). Results from propensity score weighted Cox model also showed comparable risks in these two treatment groups (HR=0.88, 95% CI: 0.45 to 1.71; P=0.700), following confirmation of successful balance of baseline covariates between groups in the weighted sample (SMDs<0.10). The HRs remained close to 1 during the sequential covariate adjustment process (ranging from 0.86-1.18 across different models; **Figure 2**). No violation of the proportional hazards assumption were detected in any model (P>0.10).

**Figure 2.**
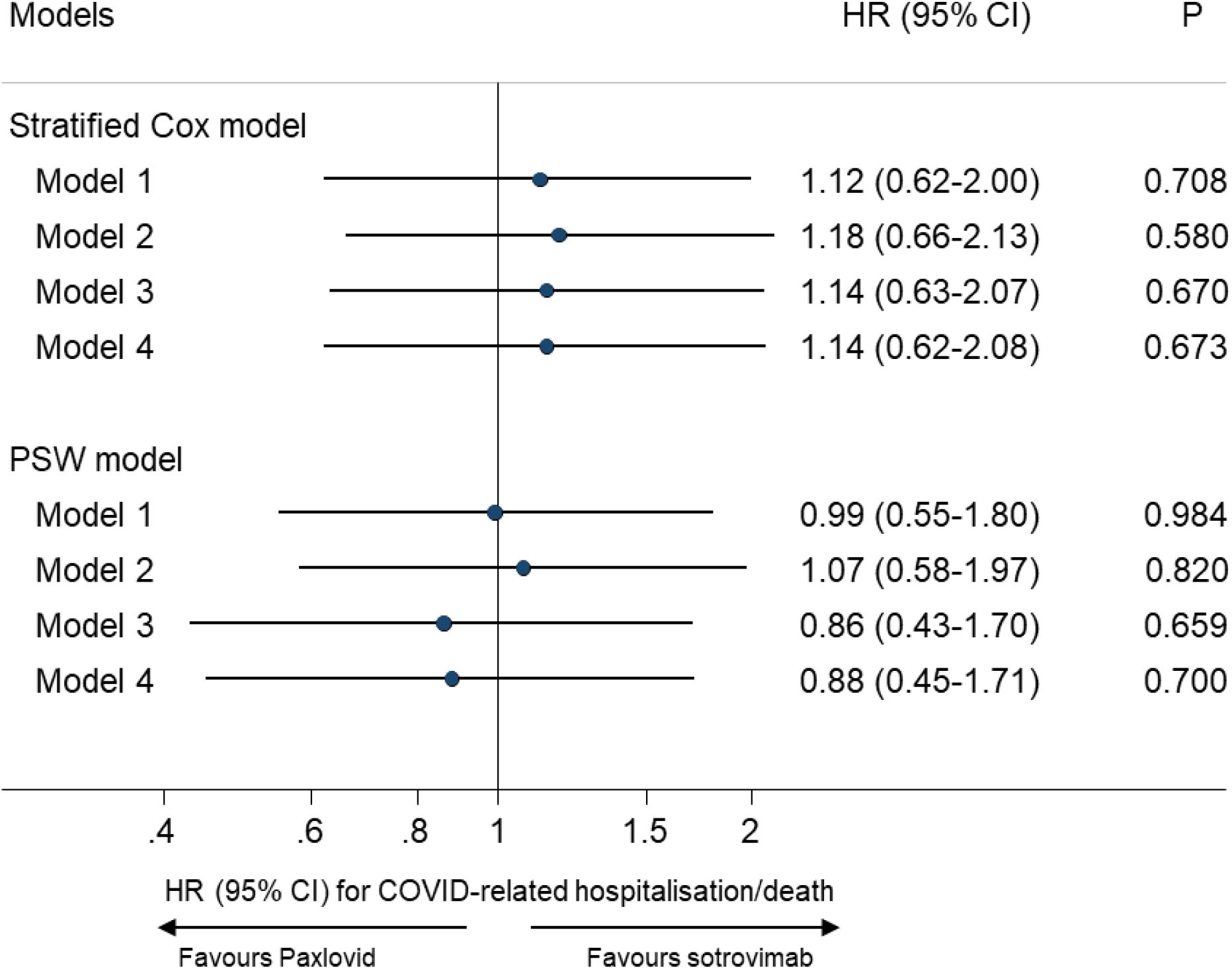
Comparing risk of 28-day COVID-19 related hospitalisation/death between Paxlovid vs. sotrovimab. Note: PSW=propensity score weighting; HR=hazard ratio; CI=confidence interval. Model 1 adjusted for age and sex; Model 2 additional adjusted for high risk cohort categories; Model 3 further adjusted for ethnicity, IMD quintiles, vaccination status, calendar date; and Model 4 additionally adjusted for BMI category, diabetes, hypertension, chronic cardiac and respiratory diseases.

For the secondary outcomes, the analysis of 60-day COVID-19 related hospitalisations/deaths also showed no evidence of difference between the treatment groups (HRs ranging from 1.01-1.33 across models; P>0.05; **Table 2**). For all-cause hospitalisations/deaths, 220 cases (2.87%) were observed during the 28 days of follow-up after treatment initiation (123 (2.55%) in the Paxlovid group and 97 (3.41%) in the sotrovimab group); results of Cox regression showed no substantial difference between the two treatment groups (HRs ranging from 0.75-0.91 across models; **Table 2**).

**Table 2.** Comparing risks of secondary outcomes between Paxlovid vs. sotrovimab.

| Outcomes | N/Events | Stratified Cox model |  | PSW analysis |  |
| --- | --- | --- | --- | --- | --- |
|  |  | HR (95% CI) for Paxlovid | P | HR (95% CI) for Paxlovid | P |
| **28-day All-cause hospitalisation/death** | 7675/220 |  |  |  |  |
| **Model 1** |  | 0.80 (0.60-1.05) | 0.102 | 0.75 (0.57-0.99) | 0.045 |
| **Model 2** |  | 0.91 (0.69-1.20) | 0.521 | 0.87 (0.66-1.15) | 0.341 |
| **Model 3** |  | 0.88 (0.66-1.17) | 0.372 | 0.81 (0.60-1.09) | 0.163 |
| **Model 4** |  | 0.89 (0.67-1.18) | 0.412 | 0.84 (0.63-1.13) | 0.248 |
| **60-day COVID-19 related hospitalisation/death\*** | 7176/62 |  |  |  |  |
| **Model 1** |  | 1.22 (0.70-2.11) | 0.479 | 1.15 (0.66-2.00) | 0.619 |
| **Model 2** |  | 1.33 (0.77-2.30) | 0.308 | 1.25 (0.71-2.20) | 0.433 |
| **Model 3** |  | 1.31 (0.75-2.30) | 0.338 | 1.01 (0.54-1.91) | 0.974 |
| **Model 4** |  | 1.33 (0.75-2.34) | 0.325 | 1.04 (0.56-1.93) | 0.910 |

Note: PSW=propensity score weighting; HR=hazard ratio; CI=confidence interval. Model 1 adjusted for age and sex; Model 2 additional adjusted for high risk cohort categories; Model 3 further adjusted for ethnicity, IMD quintiles, vaccination status, calendar date; and Model 4 additionally adjusted for BMI category, diabetes, hypertension, chronic cardiac and respiratory diseases. \* For the analysis of 60-day outcome, patients treated after September 1, 2022 were excluded to avoid missing outcome events, as the latest hospitalisation and death events recorded in our data extraction were on October 31, 2022.

### Sensitivity analyses and tests for effect modification

Results of sensitivity analyses were generally consistent with the main findings (**Supplementary Table 1**). No substantial effect modification was observed for the tested covariates, except for a suggestive signal for sex and presence of diabetes. However, the number of outcome events in these subgroup analyses were small and results should be treated with caution (**Supplementary Figure 1**).

### Exploratory analysis comparing Paxlovid with molnupiravir

During the study period, 802 eligible non-hospitalised COVID-19 patients were treated with molnupiravir, with 11 cases (1.37%) of COVID-19 related hospitalisations/deaths observed within the 28 days follow-up (≤5 deaths). Compared to the Paxlovid group, the molnupiravir group was older (57.6 vs. 53.5 years), had a higher proportion of Down syndrome, diabetes, hypertension, chronic heart diseases and chronic respiratory diseases, and a lower proportion of rare neurological conditions (**Supplementary Table 2**). There were also some geographic variations in the prescription of these two drugs and greater use of molnupiravir earlier during the study period.

Cox regression for 28-day COVID-19 related hospitalisation/death showed varied results, with strong evidence favouring Paxlovid in the propensity score weighted Cox model (HR=0.26, 95% CI: 0.10 to 0.65; P=0.004) but not in the stratified Cox model with conventional adjustment for covariates (HR=0.61, 95% CI: 0.29 to 1.29; P=0.198; **Table 3**). To explore the cause of these discrepancies we inspected the propensity scores and found that, unlike the situation in the Paxlovid vs. sotrovimab analysis, the propensity score distributions differed between Paxlovid and molnupiravir group, with a much smaller range of overlap (**Supplementary Figure 2**). Further analysis with trimming [13] showed that, after excluding both Paxlovid and molnupiravir users below the first percentile of PS in Paxlovid group and above the 99th percentile of PS in molnupiravir group, the association became weaker (N=4788, HR=0.39, 95% CI: 0.15 to 1.00). A more strict trimming threshold (using 5th and 95th percentiles respectively) yielded an estimate close to that in the stratified Cox model (N=3551, HR=0.53, 95% CI: 0.22 to 1.27).

**Table 3.** Comparing risks of outcome events between Paxlovid vs. molnupiravir.

| Outcomes | N/Events | Stratified Cox model |  | PSW analysis |  |
| --- | --- | --- | --- | --- | --- |
|  |  | HR (95% CI) for Paxlovid | P | HR (95% CI) for Paxlovid | P |
| **28-day COVID-19 related hospitalisation/death** | 5638/44 |  |  |  |  |
| **Model 1** |  | 0.63 (0.31-1.31) | 0.216 | 0.25 (0.10-0.62) | 0.002 |
| **Model 2** |  | 0.61 (0.29-1.25) | 0.177 | 0.24 (0.10-0.59) | 0.002 |
| **Model 3** |  | 0.62 (0.30-1.30) | 0.203 | 0.25 (0.10-0.63) | 0.003 |
| **Model 4** |  | 0.61 (0.29-1.29) | 0.198 | 0.26 (0.10-0.65) | 0.004 |
| **28-day All-cause hospitalisation/death** | 5634/156 |  |  |  |  |
| **Model 1** |  | 0.72 (0.48-1.08) | 0.112 | 0.55 (0.31-0.97) | 0.038 |
| **Model 2** |  | 0.70 (0.47-1.05) | 0.084 | 0.53 (0.30-0.95) | 0.032 |
| **Model 3** |  | 0.71 (0.47-1.06) | 0.097 | 0.57 (0.32-1.01) | 0.056 |
| **Model 4** |  | 0.72 (0.48-1.09) | 0.118 | 0.61 (0.34-1.12) | 0.110 |

Note: PSW=propensity score weighting; HR=hazard ratio; CI=confidence interval. Model 1 adjusted for age and sex; Model 2 additional adjusted for high risk cohort categories; Model 3 further adjusted for ethnicity, IMD quintiles, vaccination status, calendar date; and Model 4 additionally adjusted for BMI category, diabetes, hypertension, chronic cardiac and respiratory diseases.

Among the molnupiravir users, 33 cases (4.11%) of all-cause hospitalisations/deaths were observed within 28 days of follow-up (≤5 deaths). There was some, but weaker, evidence of difference between those treated with Paxlovid and molnupiravir in the risk of 28-day all-cause hospitalisation/death (HR=0.61, 95% CI: 0.34 to 1.12 in propensity score weighted Cox model and 0.72, 95% CI: 0.48 to 1.09 in stratified Cox model although confidence intervals were below 1 in earlier adjustment; **Table 3**). Similarly to COVID-19 related outcomes, we observed attenuated association in the propensity score weighted Cox model after trimming [13] (1/99th percentile: HR=0.70, 95% CI: 0.40 to 1.23; 5/95th percentile: HR=0.70, 95% CI: 0.42 to 1.16).

## Discussion

### Summary

This is one of the largest observational studies to date on the comparative effectiveness of Paxlovid versus alternative antiviral medications between February and October 2022, covering the Omicron BA.2- and BA.5-dominant periods in the UK [14,15]. Our analysis with near real-time data shows that treatment with Paxlovid is associated with a similar risk of severe outcomes from COVID-19 infection as with sotrovimab. The findings remained robust in propensity score weighting analysis and other sensitivity analysis. The exploratory analyses using molnupiravir users as the comparison group showed some evidence in favour of Paxlovid regarding the COVID-19 related outcomes, although the strength of evidence is limited and more uncertain than the main analyses due to low numbers of Paxlovid-eligible molnupiravir users, and larger differences in comparability between users of different drugs at baseline.

### Strengths and weaknesses

The key strengths of the OpenSAFELY platform are the scale, level of detail and completeness of the underlying primary care EHR data and the linkage to multiple COVID-19 relevant national databases with near real-time data update [9,16]. We used a range of analytic methods to examine robustness of results, and were able to carry out extensive adjustments for confounding and exclusions for contraindications given the availability of granular multisource real-world data. As well as availability of treatment to the population regardless of ability to pay, the uniqueness of the UK data is that the administration of COVID-19 medications in the community, is operated by CMDUs that were launched specifically for COVID-19 treatment, following the national prescription guidelines and clear eligibility criteria for treatment [2]. Therefore, our study population are well-characterised with high quality exposure data based on treatment records in the central system.

Several limitations of this study need to be considered. Despite the granular data on underlying health status, the possibility of residual confounding cannot be ruled out, in particular related to severity of COVID-19 symptoms or other unmeasured features (such as level of immunosuppression) that may have influenced clinician’s choice of therapy at assessment. This is more evident for the exploratory analysis of Paxlovid vs. molnupiravir, where the two drugs had no clinical equipoise (first-line vs. third-line) [2]. In addition, there is limited power especially for the analysis of Paxlovid vs. molnupiravir (with only 802 patients and 11 events). We used the primary care data to detect drugs which contraindicate use of Paxlovid [11], and thus could have missed prescriptions outside of the GP record (secondary care/’hospital at home’) leading to misclassification of eligibility to receive Paxlovid among sotrovimab and molnupiravir users. Finally, the patients included in this study are assumed to be only those who met the eligibility criteria made by NHS England and had no contraindications for Paxlovid [2], thus limiting further generalisation of our findings to other patient groups.

### Findings in Context

The randomised clinical trials of nMAbs and antivirals were conducted during periods where different variants of SARS-CoV-2 were circulating, and in often non-vaccinated populations, making their relevance to contemporary situations limited. Comparative trials between different treatments have not been conducted, making clinical decisions about treatment for patients eligible to receive any therapy difficult. Both the EPIC-HR trial [3] for Paxlovid and the COMET-ICE trial [4] for sotrovimab showed evidence of benefit compared to placebo (relative risk=0.12 and 0.21, respectively). However, a large-scale pragmatic study of molnupiravir, the UK PANORAMIC trial [17], showed that molnupiravir did not reduce risk of hospitalisations/deaths among high-risk vaccinated adults with COVID-19 in the community (25,000 participants, adjusted odds ratio=1.06, 95% Bayesian credible interval: 0.80 to 1.40). The PANORAMIC trial for Paxlovid versus usual care is still ongoing.

A recent literature review shows that a growing body of real-world evidence supports the efficacy of Paxlovid among vaccinated adult patients in the Omicron era [18], though most of those observational studies were conducted during the BA.1/BA.2 wave. A large-scale cohort study in Israel showed that among high-risk outpatients with COVID-19 aged ≥65 years, Paxlovid users had substantially lower risks of hospitalisation due to COVID-19 (adjusted hazard ratio=0.27, 95% CI: 0.15 to 0.49) and death due to COVID-19 (adjusted hazard ratio=0.21, 95% CI: 0.05 to 0.82) than untreated patients; whereas no association was observed in patients aged <65 years [5]. A large-scale observational study of non-hospitalised COVID-19 patients in Hong Kong SAR showed that Paxlovid use was associated with lower risks of all-cause mortality (HR=0.34, 95% CI: 0.22 to 0.52) and hospital admission due to COVID-19 (HR=0.76, 95% CI: 0.67 to 0.86) than non-use, with no effect modification by age [6]. In contrast, molnupiravir use was associated with lower risk of death (HR=0.76, 95% CI: 0.61 to 0.95) but not risk of hospitalisation (0.98, 95% CI: 0.89 to 1.06) than non-use [6]. Another cohort study in the US population showed that non-hospitalised adult patients who were prescribed Paxlovid had a lower risk of all-cause hospitalisation or death than non-users (adjusted risk ratio=0.56, 95% CI: 0.42 to 0.75) [19]. The evidence for the safety of Paxovid in routine clinical use, however, is still limited, and the concerns of viral rebound and recurrence of COVID-19 symptoms following Paxlovid treatment need further investigation [18].

### Policy Implications and Interpretation

While randomised clinical trials are rightly considered a gold standard for studying drug effectiveness, rapidly evolving SARS-CoV-2 variants can mean results of COVID-19 therapeutics trials may be quickly outdated. Additionally, there can remain areas of uncertainty where clinical trials have not been conducted such as comparative effectiveness between different therapeutic agents or for populations underrepresented in clinical trials. In vitro data can provide conclusive evidence of loss of drug effect but where different variants are circulating, or where in vitro data shows intermediate effects, the benefits of treatment in a whole population may be uncertain [20]. We believe that careful analysis of routinely-collected healthcare data can provide rapid analysis of drug effectiveness and safety, within the whole population and can provide critical information which can be used alongside in vitro data to support future decision-making [16,21].

## Summary

In routine care of non-hospitalised high-risk adult patients with COVID-19 in England, we observed no substantial difference in the risk of severe COVID-19 outcomes between those who received Paxlovid and sotrovimab during a period of Omicron BA.2 and BA.5 dominance.

## Data Availability

All data were linked, stored and analysed securely within the OpenSAFELY platform https://opensafely.org/. All code is shared openly for review and re-use under MIT open license. Detailed pseudonymised patient data is potentially re-identifiable and therefore not shared. We rapidly delivered the OpenSAFELY data analysis platform without prior funding to deliver timely analyses on urgent research questions in the context of the global Covid-19 health emergency: now that the platform is established we are developing a formal process for external users to request access in collaboration with NHS England; details of this process are available at OpenSAFELY.org.

## Administrative

## Acknowledgements

We are very grateful for all the support received from the TPP Technical Operations team throughout this work, and for generous assistance from the information governance and database teams at NHS England and the NHS England Transformation Directorate.

## Information governance and ethical approval

NHS England is the data controller; TPP is the data processor; and the researchers on OpenSAFELY are acting with the approval of NHS England. This implementation of OpenSAFELY is hosted within the TPP environment which is accredited to the ISO 27001 information security standard and is NHS IG Toolkit compliant; patient data has been pseudonymised for analysis and linkage using industry standard cryptographic hashing techniques; all pseudonymised datasets transmitted for linkage onto OpenSAFELY are encrypted; access to the platform is via a virtual private network (VPN) connection, restricted to a small group of researchers; the researchers hold contracts with NHS England and only access the platform to initiate database queries and statistical models; all database activity is logged; only aggregate statistical outputs leave the platform environment following best practice for anonymisation of results such as statistical disclosure control for low cell counts. The OpenSAFELY research platform adheres to the obligations of the UK General Data Protection Regulation (GDPR) and the Data Protection Act 2018. In March 2020, the Secretary of State for Health and Social Care used powers under the UK Health Service (Control of Patient Information) Regulations 2002 (COPI) to require organisations to process confidential patient information for the purposes of protecting public health, providing healthcare services to the public and monitoring and managing the COVID-19 outbreak and incidents of exposure; this sets aside the requirement for patient consent. Taken together, these provide the legal bases to link patient datasets on the OpenSAFELY platform. GP practices, from which the primary care data are obtained, are required to share relevant health information to support the public health response to the pandemic, and have been informed of the OpenSAFELY analytics platform.

This study was approved by the Health Research Authority (REC reference 20/LO/0651) and by the LSHTM Ethics Board (reference 21863).

## Data access and verification

Access to the underlying identifiable and potentially re-identifiable pseudonymised electronic health record data is tightly governed by various legislative and regulatory frameworks, and restricted by best practice. The data in OpenSAFELY is drawn from General Practice data across England where TPP is the Data Processor. TPP developers (CB, JC, JP, FH, and SH) initiate an automated process to create pseudonymised records in the core OpenSAFELY database, which are copies of key structured data tables in the identifiable records. These are linked onto key external data resources that have also been pseudonymised via SHA-512 one-way hashing of NHS numbers using a shared salt. Bennett Institute for Applied Data Science developers and PIs (BG, CEM, SB, AJW, KW, WJH, HJC, DE, PI, SD, GH, BBC, RMS, ID, TW, TO, SM, CLS, LB,KB, EJW and CTR) holding contracts with NHS England have access to the OpenSAFELY pseudonymised data tables as needed to develop the OpenSAFELY tools. These tools in turn enable researchers with OpenSAFELY Data Access Agreements to write and execute code for data management and data analysis without direct access to the underlying raw pseudonymised patient data, and to review the outputs of this code. All code for the full data management pipeline—from raw data to completed results for this analysis—and for the OpenSAFELY platform as a whole is available for review at github.com/OpenSAFELY.

## Guarantor

LAT/BZ are guarantors.

## References

[1] NHS. Neutralising monoclonal antibodies (nMABs) or antivirals for non-hospitalised patients with COVID-19. 2021. https://www.cas.mhra.gov.uk/ViewandAcknowledgment/ViewAlert.aspx?AlertID=103186

[2] NHS. Antivirals or Neutralising Antibodies for Non-Hospitalised Patients with COVID-19. 2022. https://www.cas.mhra.gov.uk/ViewandAcknowledgment/ViewAlert.aspx?AlertID=103208

[3] Hammond J, Leister-Tebbe H, Gardner A, et al. Oral Nirmatrelvir for High-Risk, Nonhospitalized Adults with Covid-19. N Engl J Med. 2022;386(15):1397–1408. doi:10.1056/NEJMoa2118542

[4] Gupta A, Gonzalez-Rojas Y, Juarez E, et al. Early Treatment for Covid-19 with SARS-CoV-2 Neutralizing Antibody Sotrovimab. N Engl J Med. 2021;385(21):1941–1950. doi:10.1056/NEJMoa2107934

[5] Arbel R, Wolff Sagy Y, Hoshen M, et al. Nirmatrelvir Use and Severe Covid-19 Outcomes during the Omicron Surge. N Engl J Med. 2022;387(9):790–798.

[6] Wong CKH, Au ICH, Lau KTK, Lau EHY, Cowling BJ, Leung GM. Real-world effectiveness of molnupiravir and nirmatrelvir plus ritonavir against mortality, hospitalisation, and in-hospital outcomes among community-dwelling, ambulatory patients with confirmed SARS-CoV-2 infection during the omicron wave in Hong Kong: an observational study. Lancet. 2022;400(10359):1213–1222.

[7] Agarwal A, Rochwerg B, Lamontagne F, et al. A living WHO guideline on drugs for covid-19. BMJ. 2020;370:m3379.

[8] NHS. Interim Clinical Commissioning Policy: Treatments for non-hospitalised patients with COVID-19. 2022. https://www.england.nhs.uk/coronavirus/wp-content/uploads/sites/52/2022/11/C1710-interim-clinical-commissioning-policy-treatments-for-non-hospitalised-patients-with-covid-19-nov-22.pdf

[9] Green ACA, Curtis HJ, Higgins R, et al. Trends, variation, and clinical characteristics of recipients of antiviral drugs and neutralising monoclonal antibodies for covid-19 in community settings: retrospective, descriptive cohort study of 23.4 million people in OpenSAFELY. BMJ Medicine. 2023;2:e000276.

[10] Jayk Bernal A, Gomes da Silva MM, Musungaie DB, et al. Molnupiravir for Oral Treatment of Covid-19 in Nonhospitalized Patients. N Engl J Med. 2022;386(6):509–520.

[11] Specialist Pharmacy Services. PF-07321332 [Nirmatrelvir] plus ritonavir (Paxlovid). 2022. https://web.archive.org/web/20220208181734/https://www.sps.nhs.uk/wp-content/uploads/2022/01/Paxlovid-SPS-20220202.pdf

[12] Stürmer T, Webster-Clark M, Lund JL, et al. Propensity Score Weighting and Trimming Strategies for Reducing Variance and Bias of Treatment Effect Estimates: A Simulation Study. Am J Epidemiol. 2021;190(8):1659–1670.

[13] Stürmer T, Rothman KJ, Avorn J, Glynn RJ. Treatment effects in the presence of unmeasured confounding: dealing with observations in the tails of the propensity score distribution--a simulation study. Am J Epidemiol. 2010;172(7):843–854.

[14] UK Health Security Agency. SARS-CoV-2 variants of concern and variants under investigation in England: technical briefing 48. 2022. https://assets.publishing.service.gov.uk/government/uploads/system/uploads/attachment_data/file/1120304/technical-briefing-48-25-november-2022-final.pdf

[15] Wellcome Sanger Institute. COVID–19 Genomic Surveillance. 2023. https://covid19.sanger.ac.uk/lineages/raw?latitude=52.208076&longitude=-4.847882&zoom=7.740295

[16] Zheng B, Green ACA, Tazare J, et al. Comparative effectiveness of sotrovimab and molnupiravir for prevention of severe covid-19 outcomes in patients in the community: observational cohort study with the OpenSAFELY platform. BMJ. 2022;379:e071932.

[17] Butler CC, Hobbs FDR, Gbinigie OA, et al. Molnupiravir plus usual care versus usual care alone as early treatment for adults with COVID-19 at increased risk of adverse outcomes (PANORAMIC): an open-label, platform-adaptive randomised controlled trial. Lancet. 2022;S0140-6736(22)02597-1.

[18] Akinosoglou K, Schinas G, Gogos C. Oral Antiviral Treatment for COVID-19: A Comprehensive Review on Nirmatrelvir/Ritonavir. Viruses. 2022;14(11):2540.

[19] Dryden-Peterson S, Kim A, Kim AY, et al. Nirmatrelvir Plus Ritonavir for Early COVID-19 in a Large U.S. Health System: A Population-Based Cohort Study. Ann Intern Med. 2023;176:77–84.

[20] Wu MY, Carr EJ, Harvey R, et al. WHO’s Therapeutics and COVID-19 Living Guideline on mAbs needs to be reassessed. Lancet. 2022;S0140-6736(22)01938-9.

[21] The OpenSAFELY Collaborative. Comparative effectiveness of sotrovimab and molnupiravir for preventing severe COVID-19 outcomes in non-hospitalised patients on kidney replacement therapy: observational cohort study using the OpenSAFELY-UKRR linked platform and SRR database. medRxiv. 2022; doi: https://doi.org/10.1101/2022.12.02.22283049

