## Supplementary Table 1 for "Comparative effectiveness of Paxlovid versus sotrovimab and molnupiravir for preventing severe COVID-19 outcomes in non-hospitalised patients: observational cohort study using the OpenSAFELY platform"

### Supplement

#### Supplementary Table 1. Sensitivity analysis of 28-day COVID-19 related hospitalisation/death between Paxlovid vs. sotrovimab.

| Sensitivity analysis | N | Events | HR (95% CI) for Paxlovid (ref=sotrovimab) | P |
|---|---|---|---|---|
| additionally adjusting for days between test positive and treatment initiation, and months between last vaccination date and treatment initiation | 7683 | 52 | 1.16 (0.63-2.14) | 0.63 |
| using restricted cubic splines for age | 7683 | 52 | 1.15 (0.63-2.10) | 0.656 |
| additionally adjusting for rural-urban, comorbidity, housebound | 7683 | 52 | 1.18 (0.64-2.17) | 0.592 |
| excluding patients with treatment records of both sotrovimab and molnupiravir, or with treatment records of any other therapies | 7650 | redacted | 1.14 (0.62-2.09) | 0.669 |
| excluding patients who did not have a positive SARS-CoV-2 test record before treatment or initiated treatment after 5 days since positive SARS-CoV-2 test | 6715 | 39 | 0.77 (0.38-1.53) | 0.454 |
| create a 1-day lag in the follow-up start date | 7660 | 44 | 1.15 (0.60-2.22) | 0.676 |
| create a 2-day lag in the follow-up start date | 7647 | 37 | 1.31 (0.63-2.73) | 0.465 |
| multiple imputation for covariates | 7683 | 52 | 1.14 (0.62-2.09) | 0.669 |
| Complete-case analysis | 6562 | 46 | 1.24 (0.65-2.38) | 0.514 |
| not excluding contraindications, adjusting for them as covariates | 16085 | 157 | 0.80 (0.55-1.17) | 0.243 |
| excluding contraindicated drug within 1-year before baseline | 7070 | 45 | 1.20 (0.62-2.31) | 0.59 |
| excluding contraindicated drug within 90 days before baseline | 8168 | 53 | 1.23 (0.68-2.25) | 0.495 |

**Supplementary Table 2. Baseline characteristics of patients receiving Paxlovid or molnupiravir.**

| Characteristics | Paxlovid group | Molnupiravir group |
|---|---|---|
| N | 4836 | 802 |
| Age (year), mean (SD) | 53.5 (14.8) | 57.6 (16.0) |
| Female, n (%) | 3092 (63.9) | 486 (60.6) |
| White, n (%) | 4422 (93.0) | 746 (95.0) |
| IMD quintile, n (%) | | |
| 1 (most deprived) | 467 (10.0) | 73 (9.2) |
| 2 | 723 (15.4) | 157 (19.9) |
| 3 | 1048 (22.3) | 202 (25.6) |
| 4 | 1195 (25.5) | 188 (23.8) |
| 5 (least deprived) | 1262 (26.9) | 170 (21.5) |
| Region (NHS), n (%) | | |
| East | 1180 (24.4) | 372 (46.4) |
| London | 219 (4.5) | 18 (2.2) |
| East Midlands | 1129 (23.4) | 29 (3.6) |
| West Midlands | 61 (1.3) | 8 (1.0) |
| North East | 125 (2.6) | 6 (0.7) |
| North West | 487 (10.1) | 25 (3.1) |
| South East | 474 (9.8) | 107 (3.3) |
| South West | 535 (11.1) | 162 (20.2) |
| Yorkshire | 626 (12.9) | 75 (9.4) |
| High risk cohorts, n (%) | | |
| Down syndrome | 204 (4.2) | 48 (6.0) |
| Solid cancer | 658 (13.6) | 123 (15.3) |
| Haematological disease | 729 (15.1) | 142 (17.7) |
| Immune-mediated inflammatory diseases | 1846 (38.2) | 324 (40.4) |
| Immunosuppression | 525 (10.9) | 88 (11.0) |

| Characteristics | Paxlovid group | Molnupiravir group |
|---|---|---|
| HIV/AIDS | 14 (0.3) | ≤5 |
| Rare neurological disease | 1290 (26.7) | 146 (18.2) |
| BMI (kg/m$^2$), mean (SD) | 28.1 (6.5) | 28.4 (6.6) |
| Comorbidities, n (%) | | |
| Diabetes | 603 (12.5) | 144 (18.0) |
| Chronic cardiac disease | 271 (5.6) | 89 (11.1) |
| Hypertension | 1121 (23.2) | 276 (34.4) |
| Chronic respiratory disease | 729 (15.1) | 157 (19.6) |
| Vaccination status, n (%) | | |
| None | 74 (1.5) | Combined: 17 (2.1) |
| One vaccination | 74 (1.5) | |
| Two vaccinations | 192 (4.0) | 38 (4.7) |
| Three vaccinations | 2449 (50.6) | 402 (50.1) |
| Four or more | 2047 (42.3) | 345 (43.0) |
| Days between test positive and treatment, median (IQR) | 1 (1-2) | 2 (1-2) |
| Weeks between campaign start and treatment, median (IQR) | 23 (16-30) | 18 (14-30) |

Note: IMD, BMI, ethnicity and positive test date had 153, 605, 100 and 709 missing values, respectively.

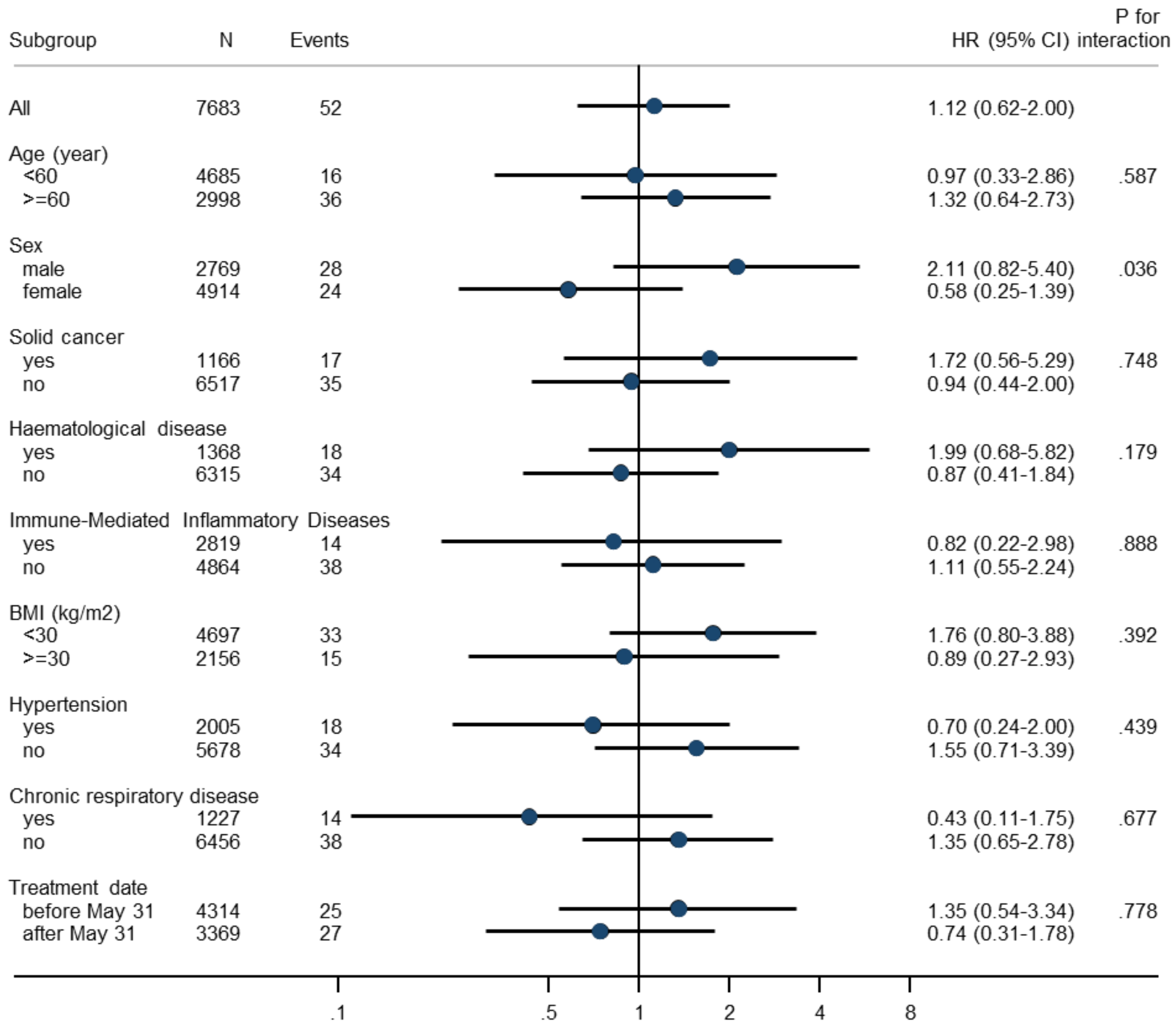


**Supplementary Figure 1. Subgroup analysis of Paxlovid vs. sotrovimab in association with risk of 28-day COVID-19 related hospitalisation/death.**

Note: HR=hazard ratio; CI=confidence interval; BMI=body mass index. Subgroup analyses were based on the fully-adjusted stratified Cox model (Model 4). P for interaction between drug group and each of the following variables (before Bonferroni correction) was: rare neurological conditions (0.078), diabetes (0.046), chronic cardiac diseases (0.723), days between test positive and treatment initiation (0.367), and White ethnicity (0.956); no analyses within each level of these variables were done because of lack of sample size or outcome events within the subset of population.

A.

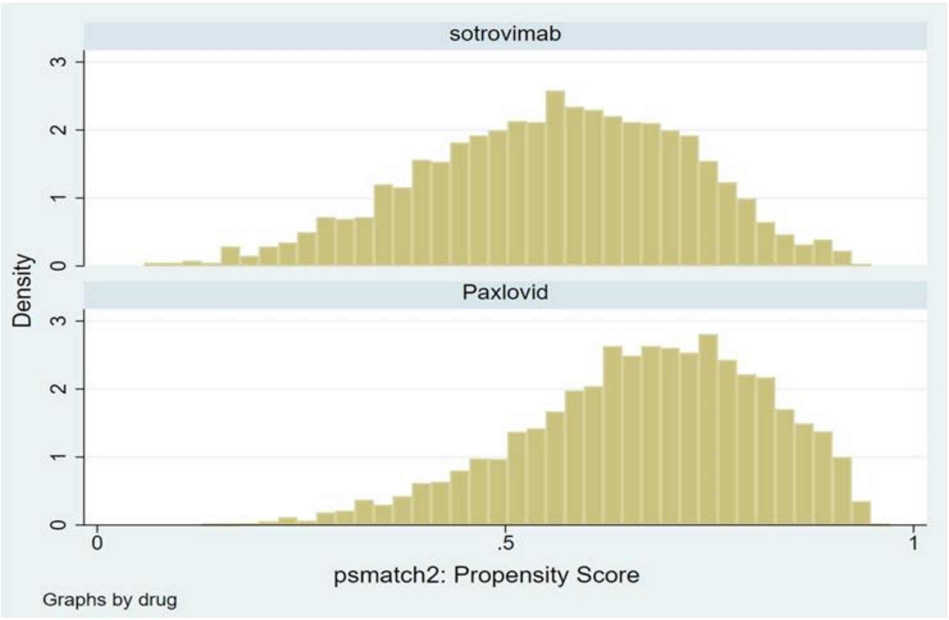


B.

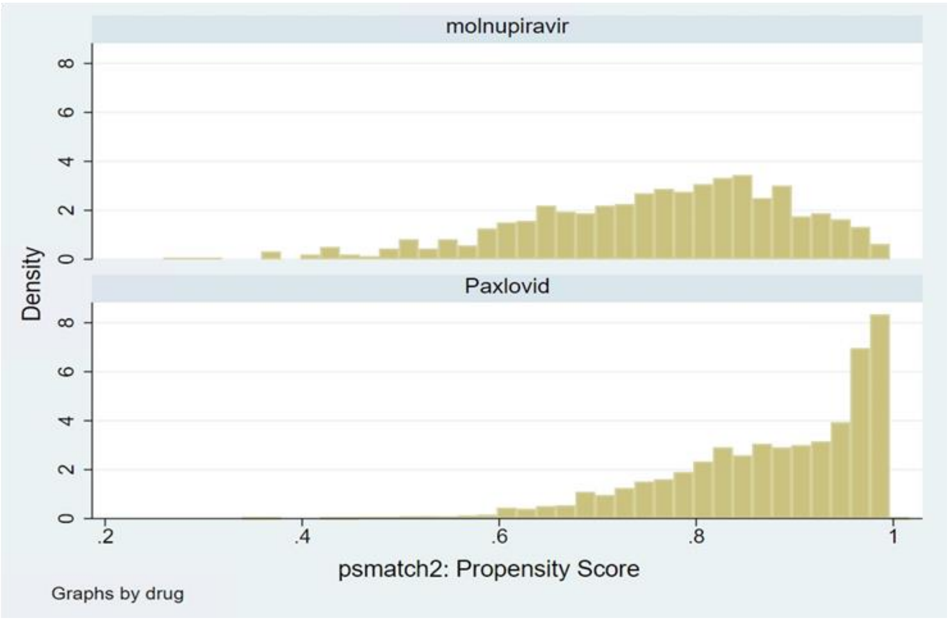


**Supplementary Figure 2. Histograms of propensity scores in Paxlovid vs. sotrovimab (A) and Paxlovid vs. molnupiravir analyses (B).**
